# Could SARS-CoV-2 be transmitted via speech droplets?

**DOI:** 10.1101/2020.04.02.20051177

**Authors:** Philip Anfinrud, Christina E. Bax, Valentyn Stadnytskyi, Adriaan Bax

## Abstract

Speaking may be a primary mode of transmission of SARS-CoV-2. Considering that reports of asymptomatic transmission account for 50-80% of COVID-19 cases and that saliva has peak viral loads at time of patient presentation, droplet emission while speaking could be a significant factor driving transmission and warrants further study. We used a planar beam of laser light passing through a dust-free enclosure to detect saliva droplets emitted while speaking. We found that saying the words ‘Stay Healthy’ generates thousands of droplets that are otherwise invisible to the naked eye. A damp homemade cloth face mask dramatically reduced droplet excretion, with none of the spoken words causing a droplet rise above the background. Our preliminary findings have important implications for pandemic mitigation efforts.

---

TO THE EDITOR:

The novel coronavirus 2019 (COVID-19) pandemic is spreading world-wide at an alarming rate. In a March 25, 2020 audio editorial, Dr. Eric Rubin noted that “Aerosols may not be the primary mode of transmission. It seems more likely that droplets are important.”^1^ We agree, yet the scientific and medical communities have been slow to study the role droplets produced during normal speech may play in the transmission of COVID-19. Considering that reports of asymptomatic transmission account for 50-80% of COVID-19 cases, droplet emission while speaking could be a significant factor driving transmission. For example, recent data^2^ show that the nasal and throat swabs of COVID-19 positive individuals contain high viral loads, with the upper respiratory viral load approaching peak levels at symptom onset. Nasal and throat viral titers were similar in the single asymptomatic patient and symptomatic patients.^3^ Similarly, Chan et al.^4^ demonstrated significant viral load in oral fluids (predominantly saliva) using a highly sensitive SARS-CoV-2 RdRp/Hel assay. Furthermore, bacterial dispersal was observed during a simulated ophthalmologic procedure but was significantly decreased by facemasks or silence.^5^

In this Letter we describe our use of laser light-scattering to sensitively detect droplet emission while speaking. Our preliminary findings have important implications for pandemic mitigation efforts.

Using a planar beam of laser light passing through a dust-free enclosure to detect saliva droplets emitted while speaking, we found that saying the words ‘Stay Healthy’ generates thousands of droplets that are otherwise invisible to the naked eye (Fig. 1). Their abundance and brightness appear to be in agreement with those previously identified by phonetics and increase with loudness.^6^ The number of droplets seen in a single frame of the video (16.6 ms duration) was as high as 360 (Fig 1A). A damp homemade cloth face mask dramatically reduced droplet excretion, with none of the spoken words causing a droplet rise above the background (Fig. 1A).

**Fig 1.**
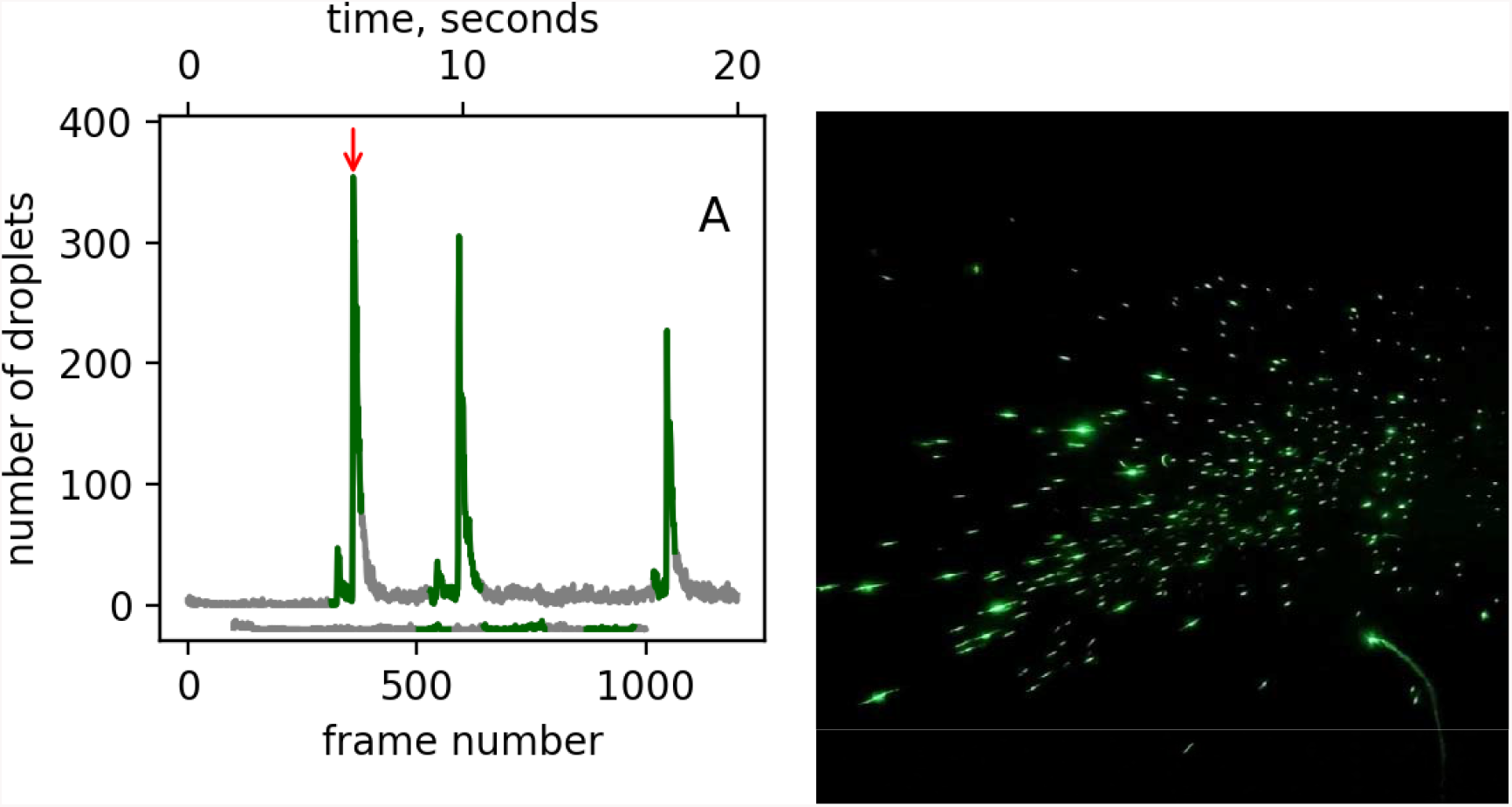
Visualizing droplet emission when speaking ‘Stay Healthy three times.’ (A) Number of droplets detected in each frame of a video acquired at a 60-Hz frame rate with and without a damp cloth face mask. Green indicates when the words were spoken. The trace offset below the major graph shows the absence of droplets during speech with mask coverage. (B) Frame # 361 from the video clip, which corresponds to the peak in the number of speech droplets detected; see red arrow in (A). The spots vary in brightness due to differences in size. The particle count after each repeated phrase remains above the background observed prior to the first repeat, suggesting that some of the speech droplets become aerosolized and linger inside the box for multiple seconds. Full video recordings are included as SI.

Droplets emitted while speaking are much smaller than those emitted when coughing or sneezing.^6^ Nonetheless they are sufficiently large to carry a variety of respiratory pathogens, including the measles virus, influenza virus, and Mycobacterium tuberculosis.^6^ Moreover, multiple studies have shown that speaking actually produces significantly *more* droplets than coughing.^6,7^ Due to their small size, specialized equipment is required to detect their abundance. The laser light-scattering method reported here detects far more droplets while speaking than previously reported with other methods.^6^

During speech in asymptomatic or healthy individuals, the majority of oral fluid is saliva with very little contribution from nasal or bronchial secretions. Speech droplets can transfer virus either through a direct pathway, by inhaling a droplet from a carrier, or from droplets landing on surfaces, followed by fomite transmission. We cannot assess the relative importance of these pathways but propose that their aggregate is key as oral fluid viral loads are already approaching peak levels by the time the patient presents,^8^ and asymptomatic transmission is common. Further studies are needed to assess the viral titer present in speech-induced droplets in asymptomatic but COVID-19 positive persons, but our results suggest that speaking can indeed be a major mode of SARS-CoV-2 transmission. Our preliminary findings therefore have vital implications for pandemic mitigation efforts: If speaking and oral fluid viral load proves to be a major mechanism of SARS-CoV-2 transmission, wearing any kind of cloth mouth cover in public by every person, as well as strict adherence to social distancing and handwashing, could significantly decrease the transmission rate and thereby contain the pandemic until a vaccine becomes available.

## Data Availability

All data is available upon request.

https://doi.org/10.5281/zenodo.3732625

## Supplementary Information

In response to the urgency of the current pandemic, our preliminary data were recorded by a rapidly repurposed optical arrangement that employed a Coherent Verdi laser operating at 2.5 W optical power and a pair of spherical (−25-mm f.l.) and cylindrical (40.6-mm f.l.) lenses to generate a light ‘sheet’ approximately 1-mm thick and 150-mm tall. This light sheet passed through slits on the sides of a cardboard box (53 cm width; 46 cm height; 62-cm depth) whose interior was painted black. The box was positioned under a HEPA filter to eliminate scattering from dust particles. When speaking through the open end of the box, speech droplets traversed approximately 50-75 mm before encountering the light sheet. An iPhone 11 Pro video camera viewed the light sheet through a 7-cm diameter hole on the opposite side of the box and recorded sound and video of light scattering events as droplets passed through this sheet. Python software was developed to analyze frame-by-frame the movie clips recorded. Video clips acquired while speaking with and without a face mask are available at https://doi.org/10.5281/zenodo.3732625.

